# Persistent immune abnormalities discriminate post-COVID syndrome from convalescence

**DOI:** 10.1101/2023.05.02.23289345

**Authors:** Julia Sbierski-Kind, Stephan Schlickeiser, Svenja Feldmann, Veronica Ober, Eva Grüner, Claire Pleimelding, Leonard Gilberg, Isabel Brand, Nikolas Weigl, Mohamed I. M. Ahmed, Gerardo Ibarra, Michael Ruzicka, Christopher Benesch, Anna Pernpruner, Elisabeth Valdinoci, Michael Hoelscher, Kristina Adorjan, Hans Christian Stubbe, Michael Pritsch, Ulrich Seybold, Johannes Bogner, Julia Roider, the Post Covid Care and KoCo19 study groups

**Author notes:** Corresponding authors., Department of Medicine IV, University Hospital, Ludwig-Maximilians-Universität München, Munich, Germany.

## Abstract

Innate lymphoid cells (ILCs) are key organizers of tissue immune responses and regulate tissue development, repair, and pathology. Persistent clinical sequelae beyond 12 weeks following acute COVID-19 disease, named post-COVID syndrome (PCS), are increasingly recognized in convalescent individuals. ILCs have been associated with the severity of COVID-19 symptoms but their role in the development of PCS remains poorly defined. Here we used multiparametric immune phenotyping, finding expanded circulating ILC precursors (ILCPs) and concurrent decreased group 2 innate lymphoid cells (ILC2s) in PCS patients compared to well-matched convalescent control groups at > 3 months after infection. Patients with PCS showed elevated expression of chemokines and cytokines associated with trafficking of immune cells (CCL19/MIP-3b, FLT3-ligand), endothelial inflammation and repair (CXCL1, EGF, RANTES, IL1RA, PDGF-AA). These results define immunological parameters associated with PCS and might help find biomarkers and disease-relevant therapeutic strategies.

## Introduction

Viral infections can result in chronic symptoms that persist in previously healthy convalescent individuals across a wide range of viral families, including Ebola virus, influenza, Epstein-Barr virus, and dengue fever^1,2^. The main symptoms are exertion intolerance, fatigue, neurocognitive and sensory impairment, sleep disturbances, flu-like symptoms, myalgia/arthralgia, and a plethora of nonspecific symptoms^3^. These post-acute infection syndromes (PAIS) are associated with autoimmunity and endothelial dysfunction, affecting both large and small vessels^3,4^; however, risk factors and the underlying pathophysiology remain largely unknown.

The COVID-19 pandemic, caused by infection with severe acute respiratory syndrome coronavirus 2 (SARS-CoV-2), has led to an increasing prevalence of convalescent patients with prolonged and persistent sequelae following acute SARS-CoV-2 infection-known as ‘long COVID’ or ‘post-COVID syndrome’^5^. The estimated prevalence of PCS ranges from 5-50%^6^, thus presenting an enormous global health burden, and can affect both patients with mild or severe forms of acute COVID-19 disease^7^. Clinical symptoms include fatigue, malaise, depression, cognitive impairment, persistent cough, dyspnoea, palpitations, and headaches^8^. While the acute phase of COVID-19 has been extensively studied, providing health care professionals with efficient treatment options, the pathogenesis of PCS remains unclear, with current hypotheses including autoimmunity, latent virus reactivation, tissue, and endothelial damage^9^.

The extreme respiratory distress in patients with acute COVID-19 is mediated primarily by immunopathology and systemic inflammation. Pathological immune signatures suggestive of T cell exhaustion, delayed bystander CD8^+^ T cell activation, and higher plasma GM-CSF and CXCL10 levels are associated with severity of the disease^10^,^11,12^. Survivors of severe COVID-19 show persistent immune abnormalities, including elevated levels of pro-inflammatory cytokines^13^. In addition to systemic inflammation, SARS-CoV-2 infects endothelial cells, causing virus-mediated apoptosis and consecutive endotheliitis and, thus, may promote endothelial damage and increased recruitment of activated immune cells into the endothelium and surrounding tissue^14^.

Dysregulated respiratory CD8^+^ T cell responses may contribute to impaired tissue conditions and development of pulmonary sequelae^15^. Recent work identified persistent immunological dysfunction in patients with post-acute sequelae of COVID-19, including highly activated innate immune cells and marked differences in specific circulating myeloid and lymphocyte populations^16^,^17^.

Innate lymphoid cells (ILCs) are tissue-resident effector immune cells with crucial roles in normal tissue development and remodeling^18,19^. These cells also participate in both protective and pathologic immune responses during lung tissue perturbation^20^,^21^. Several studies detected a reduction in total circulating ILCs in severe COVID-19 patients, while relative group 2 innate lymphoid cells (ILC2) levels, particularly NKGD^+^ ILC2s, were increased^22^,^23^. Although ILCs appear central to lung infection and repair, their role in PCS remains critically unexplored.

Here we used multicolor flow cytometry and multiplex cytokine assays on plasma from (1) healthy, uninfected controls (n=32, ‘HC’); (2) previously SARS-CoV-2-infected individuals in the convalescent phase without persistent symptoms (n=32, convalescent controls, ‘CC’); and (3) individuals with persistent symptoms following acute COVID-19 (n=27, post-COVID, ‘PC’) to identify specific immunological alterations, including ILCs, in PCS. Among the CC and PC groups, most participants were non-hospitalized during acute SARS-CoV-2 infection and CC and PC individuals had persistent symptoms for more than 12 weeks from the initial infection. We found expanded circulating ILC precursors (ILCPs) in PC individuals while ILC2s were decreased. Patients with persistent symptoms also displayed elevated pro-inflammatory cytokines (IL-8, IL-6), chemokines associated with trafficking of immune cells (CCL19/MIP-3b, FLT3-Ligand), and endothelial inflammation and repair (CXCL1, EGF, RANTES, PDGF-AA).

## Results and Discussion

### Clinical characteristics of study participants

Patients, enrolled in the Post-COVID-Care study at the LMU University Hospital Munich, presented with persistent symptoms for more than 12 weeks following acute SARS-CoV-2 infection (PC group; n=27) and were compared to convalescent patients without persistent symptoms (CC group; n=32) and healthy controls (HC group; n=32), enrolled in the KoCo19-index study (**Fig. 1A**). Clinical demographics of both study cohorts are reported in **Table 1**. The PCS, convalescent, and healthy control groups were well-matched in sex (67% female PC; 56% female CC; 53% female HC; Chi-square: 1.185, d.f. = 2), age (mean 37.15 years old PC; mean 36.09 years old CC; mean 35.91 years old HC; Kruskal-Wallis test p=0.9276), and BMI (mean BMI PC group 24.0kg/m^2^; mean BMI CC group 23.4kg/m^2^; mean BMI HC group 25.2kg/m^2^; Kruskal-Wallis test p=0.3315) (**Fig. 1B and Table 1**). Only 2 patients with COVID-19 sequelae were hospitalized during acute infection, whereas none of the convalescent study participants were hospitalized (**Fig. 1C**), reflecting that some patients experience long-term health-consequences after acute COVID-19, regardless of disease severity. For PC and CC groups, elapsed days since initial SARS-CoV-2 infection were different in median times from acute disease (113 days for PC group vs. 273 days for CC group; Mann-Whitney p= <0.0001) (**Fig. 1D**); however, initial enrolment and collection of blood for immunophenotyping took place more than 3 months after onset of COVID-19 and none of the convalescent participants reported persistent symptoms after acute disease. Acute SARS-CoV-2 infections within the PC group occurred in the period when the Omicron BA.2 variants were dominant (between January and March 2022), whereas participants of the convalescent group were confirmed to be infected with SARS-CoV-2 between March and April 2020, when parental strains drove the majority of new cases. While several risk factors, including comorbidities and virus variants, have been identified for the development of PCS^24^, clinical symptoms are similar for different SARS-CoV-2 strains, with the exception of musculoskeletal pain, where chronic burden may be lower for Omicron compared to Delta variants^25^. Consistent with numerous previous reports of PCS, the most common reported symptoms included constitutional symptoms, such as fatigue (93%) and insomnia (41%), and neurological symptoms, such as impaired alertness (74%), memory impairment (59%), and impaired speech (56%). Cardiac symptoms, including palpitations (59%), chest pain (52%), and reduced muscular strength (26%) were also a common complaint (**Fig. 1E**).

**Table 1:** Clinical and demographic characteristics of study cohorts. Data are given as numbers (percentages). BMI, body mass index. Sex, age and BMI were comparable between groups with an overall mean age of 36 years, 58% females and BMI of 24.2kg/m^2^.

|  |  | Healthy Controls (HC) | Convalescent Controls (CC) | Post Covid (PC) |
| --- | --- | --- | --- | --- |
| n |  | 32 | 32 | 27 |
| Sex | Male | 15 (46.9%) | 14 (43.8%) | 9 (33.3%) |
|  | Female | 17 (53.1%) | 18 (56.2%) | 18 (66.7%) |
| Age (years) | 20-29 | 6 (18.8%) | 6 (18.8%) | 4 (14.8%) |
|  | 30-39 | 10 (31.2%) | 12 (37.5%) | 10 (37.0%) |
|  | 40-49 | 15 (46.9%) | 12 (37.5%) | 8 (29.6%) |
|  | >49 | 1 (3.1%) | 2 (6.2%) | 5 (18.5%) |
|  | Mean | 36 | 36 | 37 |
| BMI (kg/m<sup>2</sup>) | <18,5 | 3 (9.4%) | 0 (0%) | 0 (0%) |
|  | 18,5-25 | 14 (43.8%) | 22 (68.8%) | 15 (55.6%) |
|  | 25-30 | 10 (31.2%) | 9 (28.1%) | 4 (14.8%) |
|  | >30 | 5 (13.6%) | 1 (3.1%) | 3 (11.1%) |
|  | Mean | 25.2 | 23.4 | 24 |
| Time from PCR to visit median (in days) |  |  | 273 (Min:125; Max: 318) | 113 (Min: 89; Max: 292) |

Data are given as numbers (percentages). BMI, body mass index.

**Figure 1:**
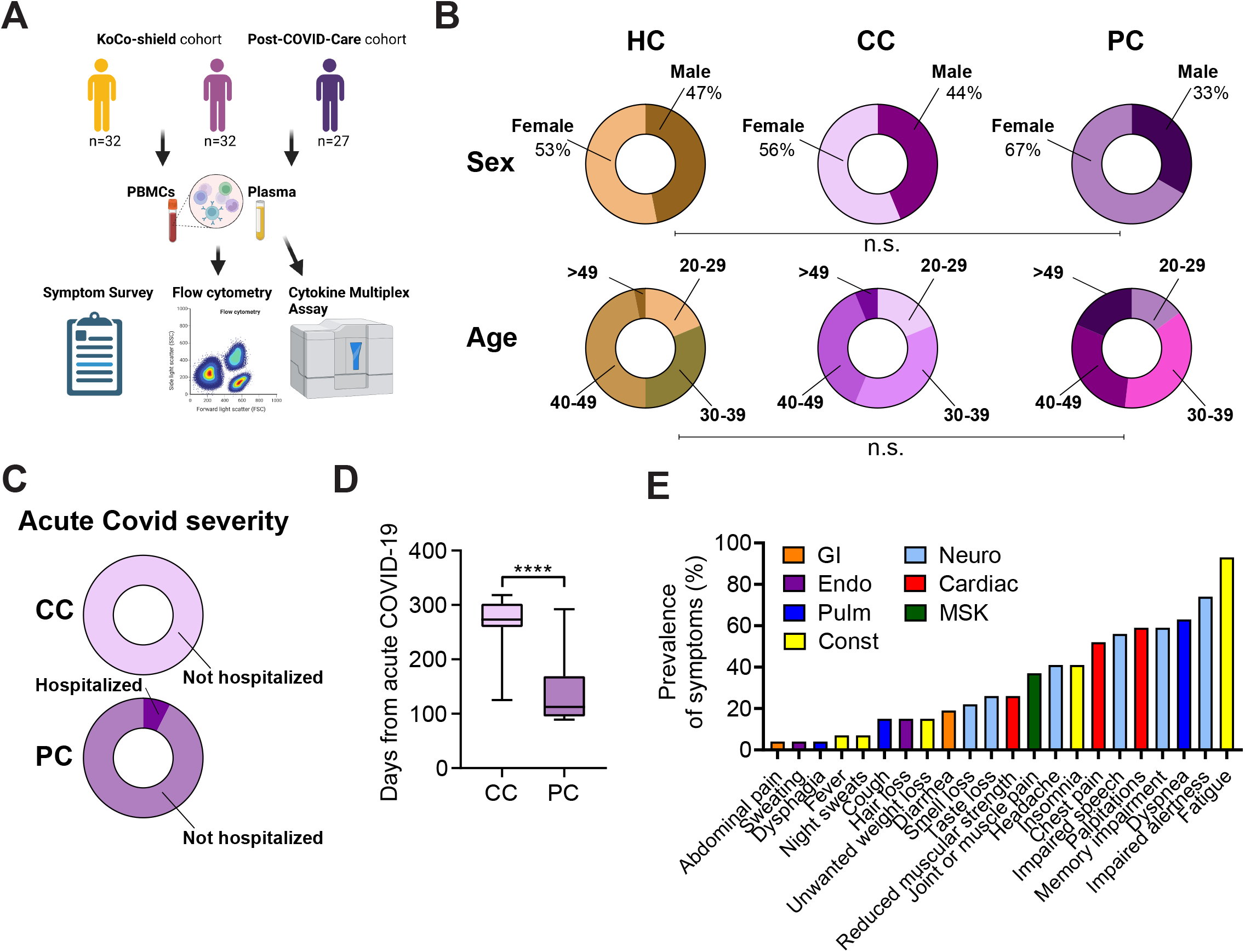
Clinical characteristics of study cohorts. (A) Overview of study cohorts and methods. The figure is partly created with BioRender.com. (B) Demographic characteristics for healthy controls with no prior SARS-CoV-2 infection (HC), convalescent SARS-CoV-2 participants without persisting symptoms (CC) and convalescent SARS-CoV-2 participants with persisting symptoms (PC) displayed as ring charts. Statistical significance is shown by capped lines as Chi-square tests for ‘Sex’ and post-hoc comparisons for ‘Age’. Characteristics are further detailed in Table 1. (C) Percentage of hospitalization during acute Covid infection for CC and PC participants displayed as ring charts. (D) Box plots showing days from first positive PCR test for CC and PC groups. Central lines indicate group means; top and bottom lines indicate minimum and maximum. Significance was assessed by Mann-Whitney-test. (E) Prevalence of top 22 self-reported symptoms in PC participants ranked from least prevalent (left) to most prevalent (right). Symptoms are colored according to physiological systems. Gastrointestinal (GI), endocrine (Endo), pulmonary (Pulm), constitutional (Const), neurological (Neuro), cardiac, and musculoskeletal (MSK). ****p ≤ 0.0001. See also Table 1.

### Pro-inflammatory cytokines and growth factors are elevated in PCS

In COVID-19 patients with severe disease, cytokine storm and uncontrolled inflammatory responses, including endothelial inflammation and associated tissue damage, are recognized as one of the driving immunopathological features that can lead to death^10^. To uncover the immunological dysregulation in PCS, we quantified 46 molecular analytes in the plasma of patients from the CC and PC groups >3 months after acute SARS-CoV-2 infection using a multiplex cytokine assay and compared them to healthy controls. Four key pro-inflammatory cytokines (interleukin-8 (IL-8), IL-6, interleukin-1 receptor antagonist (IL-1Ra) and IL-1a) were elevated in the PC group compared to the CC and HC groups (**Fig. 2A, Suppl. Fig. 1**), while no difference was observed in transforming growth factor alpha (TGF-α), IL-7, IL-5, IL-4, IL-13, IL-10, tumor necrosis factor (TNFα), interferon γ (IFN-γ) and IL-1β (**Suppl. Fig.1**). IL-8 has been previously associated with a prothrombotic neutrophil phenotype in severe COVID-19 and blocking IL-8 signaling reduced SARS-CoV-2 spike protein-induced, human ACE2-dependent pulmonary microthrombosis in mice^26^. Surprisingly, levels of IL-8 and IL-6 were lower in CC compared to HCs, whereas other pro-inflammatory cytokines were not different between these groups (**Fig. 2A, Suppl. Fig. 1**). IL-1Ra was 2.16-fold higher in the PC group compared to the HC group and 2.22-fold higher compared to the CC group; other pro-inflammatory cytokines were only slightly increased (**Fig. 2A, Suppl. Fig.1**). Several chemokines (RANTES, MIP3b, CXCL1) and growth factors (Fms-related tyrosine kinase 3 ligand (FLT-3 Ligand), epidermal growth factor (EGF), vascular endothelial growth factor (VEGF), platelet-derived growth factor A (PDGF-AA)), that could be associated with trafficking of immune cells (MIP3b, FLT3-Ligand) and endothelial inflammation (CXCL1, EGF, RANTES, PDGF-AA), CD40L and Granzyme B were also elevated in PC participants compared to both CC and HC groups (**Fig. 2A, Suppl. Fig.1**). Interestingly, Eotaxin (CCL11), monocyte chemoattractant protein 1 (MCP1), and IL-12p70 were decreased in PC patients compared to both convalescent and healthy controls (**Suppl. Fig. 1**); some of these chemokines were associated with severe cases of acute COVID-19^27^. Importantly, programmed death-ligand 1 (PD-L1) was increased in the persistent symptom group compared to both convalescent and healthy control groups, consistent with previous reports, highlighting the prognostic role of sPD-L1 in COVID-19 patients^28^ (**Suppl. Fig.1**). Although most plasma samples from the CC group were taken at month 8 after acute COVID-19 infection while samples from the PC group were taken at month 4, challenging direct comparison of persistent symptom and convalescent groups, several studies have shown that pro-inflammatory cytokines remained significantly elevated in PC patients at month 8 after acute infection^17^. Together, these data suggest persistent immune abnormalities in patients suffering from post-acute sequelae of COVID-19.

**Figure 2:**
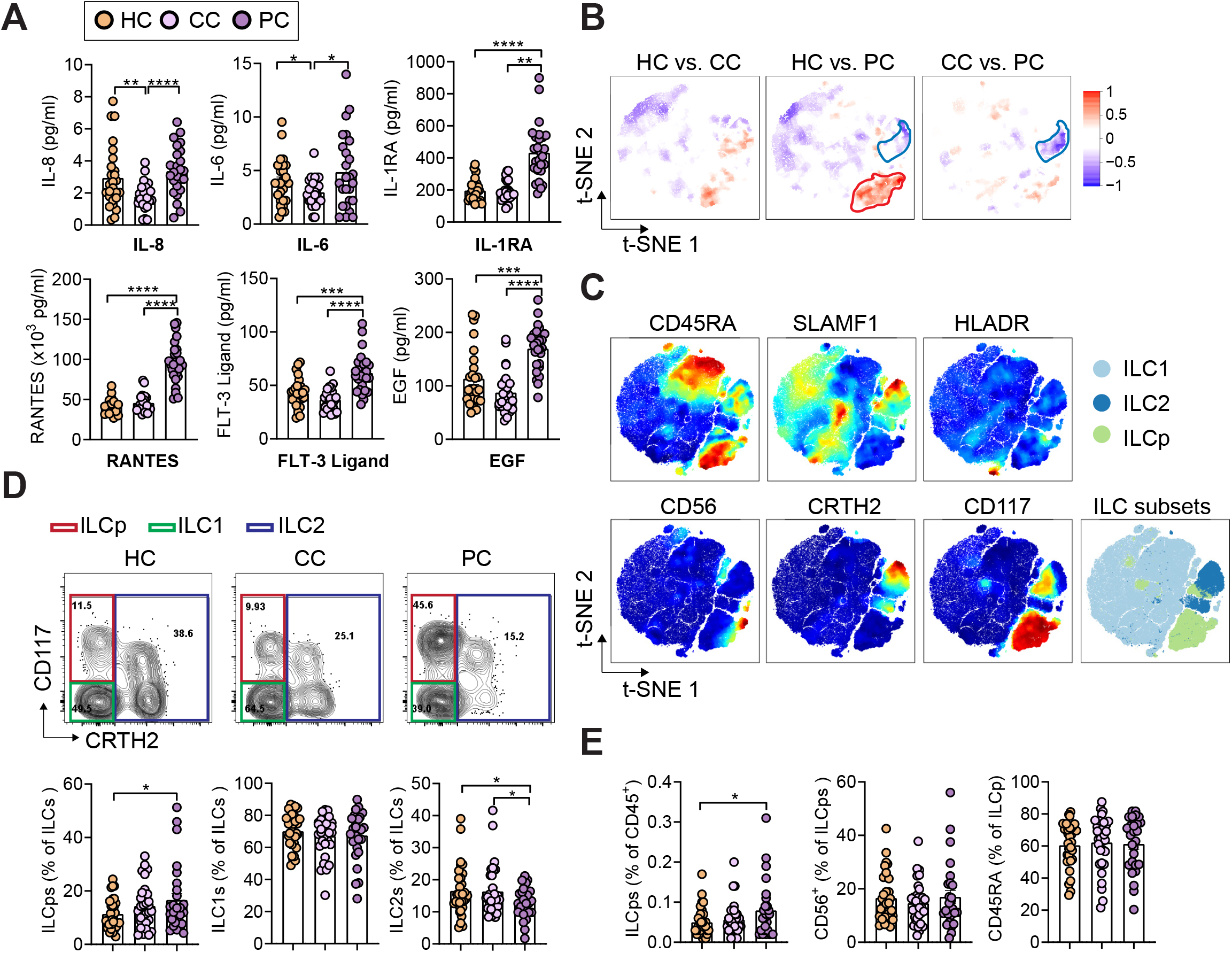
Post-COVID participants show altered cytokine expression and levels of innate lymphoid cells. (A) Multiplex assay quantification showing plasma levels of IL-8, IL-6, IL-1RA, RANTES, FLT-3 Ligand and EGF in healthy controls with no prior SARS-CoV-2 infection (HC), convalescent SARS-CoV-2 participants without persisting symptoms (CC), and convalescent SARS-CoV-2 participants with persisting symptoms (PC) at 3-10 months after acute COVID infection. (B) High-dimensionality reduction analysis of innate lymphoid cells (ILCs, gated as lymphocytes, singlets, and CD45^+^CD3^-^Lin^-^CD127^+^ cells as shown in Supplementary Figure 3) from peripheral blood mononuclear cells (PBMCs) of HC, CC, and PC groups. High-resolution group differences were visualized by calculating Cohen’s D for a given comparison across the t-SNE map. Residual plot showing differences between maps. Phenotypes within red circles were confirmed to be statistically more common in PC samples, and phenotypes within blue circles were less common in PC samples. Analysis is based on flow cytometry data from 32 HC, 32, CC, and 27 PC samples. (C) Relative expression intensities (combined HC, CC, and PC samples) of parameters used in the t-SNE analysis. (D and E) Representative flow cytometry plots (D) and quantification (D and E), showing percent innate lymphoid cell populations in HC, CC, and PC groups at 3-10 months after acute COVID infection. Bar graphs indicate mean (±SE), n=27-32 individuals per group, unpaired t-test (A, D, E), *p ≤ 0.05, **p ≤ 0.01, ***p ≤ 0.001, ****p ≤ 0.0001. See also Supplementary Figure 1.

### Circulating ILCPs are elevated in PC patients with concurrent decrease in ILC2s

To investigate circulating ILC levels via flow cytometry in PCS, convalescent and healthy controls, we used a well-established gating strategy^29^ (**Suppl. Fig.2**). Lin^-^ CD127^+^ ILC subsets were defined as CD117^-^CRTH2^-^ ILC1s, CD117^+^ ILC progenitors (ILCP)^30^, and CRTH2^+^ ILC2s. We used CD56 as a marker of activated or ILC3/NK cell-committed ILCP and CD45RA for naïve ILCP^29^. Recent work discovered CD45RA^+^ naïve-like ILCs, lacking proliferative activity, indicative of cellular quiescence^31^. To visualize multiple dimensions in simple two dimensional plots and compare flow cytometry data between groups, we used stochastic neighbor embedding analysis (**Fig. 2 B, C**). We found increased expression of the ILCP marker CD117 in PC compared to HC groups, while CRTH2 (marker for ILC2s) was decreased in PC compared to both CC and HC groups (**Fig. 2B, C**). However, the expression of proteins associated with ILC activation, CD56 (also defining NK cells with intermediate or high expression levels) and HLA-DR, was not different between groups (**Fig. 2B, C**). Next, we evaluated total numbers and frequencies of circulating ILCs and NK cells in patients with persistent symptoms after COVID-19 infection as compared to convalescent patients and healthy controls. We did not observe significant changes in total ILCs and subsequent ILC subsets (ILC2s, ILC1s, ILCPs) in PC compared to CC and HC groups (**Suppl. Fig. 3A**). However, PC patients had expanded levels of ILCPs with concurrent decreased ILC2 frequencies, while ILC1 levels remained unchanged (**Fig. 2 D, E**). The role of ILC2s in viral-induced lung pathogenesis remains controversial. Although increased levels of IL-18, IL-13, and IL-6 have been reported along with accumulation of ILC2s during acute COVID-19, increased circulating ILC2s in moderate but not severe COVID-19 patients were found in other studies^32^, consistent with their attrition by IFN-γ in type 1 (viral-induced) inflammation^20^. Thus, ILC2s might have important roles in tissue repair during viral-induced epithelial cell damage, perhaps through crosstalk with other ILC subsets.

Recent work suggested that human ILCPs can interact with endothelial cells, fostering the adhesion of other innate and adaptive immune cells by stimulating pro-inflammatory cytokine expression of adhesion molecules. This activation occurs through the tumor necrosis factor receptor- and RANK-dependent engagement of *NF-κB* pathway^33^. Nevertheless, PC patients did not show significant changes in CD45RA^+^ ILCPs, although CD56^+^ ILCPs were trending upwards, suggesting a circulating ILCP expansion without overt altered activation (**Fig. 2E**). Surprisingly, the expression of CD45RA was increased in both ILC1 and ILC2 subsets in the PC group compared to HC (**Suppl. Fig. 3B**), suggesting the increase of a quiescent local reservoir for the generation of differentiated ILCs^31^. Frequencies of HLA-DR^+^ ILC1s and the transcriptional expression of SLAMF1 within the ILC2 compartment were similar between PC, CC, and HC groups (**Suppl. Fig. 3B**). We could not find significant differences in NK cell frequencies between patients with PCS, convalescent, and healthy controls (**Suppl. Fig. 3C**). Together, these data indicate that ILCPs expand in patients with COVID-19 sequelae, without alteration of their activation state.

## Concluding remarks

Persistent sequelae following acute COVID-19 are increasingly recognized in convalescent individuals. Our exploratory analyses identified immunological differences in patients with PCS as compared to well-matched convalescent and HC individuals at > 3 months post infection. We found significant changes in circulating ILC subsets, including increased ILCPs and concurrent decreased ILC2 levels. In addition, pro-inflammatory cytokines (IL-8, IL-6), chemokines associated with trafficking of immune cells (CCL19/MIP-3b, FLT3-Ligand) and endothelial inflammation and -repair (CXCL1, EGF, RANTES, PDGF-AA) were elevated in PC participants. Although our work does not dissect how ILCPs or other activated innate and adaptive immune cells, contribute mechanistically to endothelial dysfunction in PCS, ILCP expansion along with elevated markers for endothelial inflammation in PC supports their interaction with endothelial cells; thereby facilitating enhanced inflammatory responses and endotheliitis in several organs. These findings may not only be interesting for long-term sequelae of COVID-19, but also for other viral infections that can result in PAIS in convalescent individuals. Further exploration of immunological alterations in PCS may delineate mechanisms of ILC-endothelial cell crosstalk and lead to disease-relevant targeted therapies.

## Data Availability

All data produced in the present study are available upon reasonable request to the authors.

## Abbreviations

EFG: epidermal growth factor
FLT-3 Ligand: Fms-related tyrosine kinase 3 ligand
IF-γ: interferon γ
ILC: Innate lymphoid cells
ILCP: Innate lymphoid cell precursors
ILC2s: Group 2 innate lymphoid cells
IL: interleukin
IL1RA: interleukin-1 receptor antagonist
PBMCs: Peripheral blood mononuclear cells
PAIS: Post-acute infection syndromes (PAIS)
PC: Post-Covid
PCS: Post-COVID-Syndrome
PDGF-AA: platelet-derived growth factor A
SARS-CoV-2: Severe acute respiratory syndrome coronavirus 2
TNF: tumor necrosis factor
t-SNE: t-distributed stochastic neighbor embedding
VEGF: vascular endothelial growth factor

## Data availability statement

Further information and requests for resources and reagents should be delivered to and will be fulfilled by the Lead Contact, Julia Roider.

## Ethics approval statement for human studies

<u>KoCo19 Shield</u>: The study protocol was reviewed and approved by the Institutional Review Board of the Medical Faculty at Ludwig-Maximilians-University Munich, Germany under the project number 20-692 (vote of approval dated Sept. 21st, 2020) and 20-371 (vote of approval dated May 15th, 2020). Oral and written informed consent was obtained from all study subjects. <u>PCC</u>: The study protocol was reviewed and approved by the Institutional Review Board of the Medical Faculty at Ludwig-Maximilians-University Munich, Germany under the project number 21-1165 (vote of approval dated Feb 15, 2021, amendment approved Aug. 11, 2021). Oral and written informed consent was obtained from all study subjects.

## Patient consent statement

The patients/participants provided their written informed consent to participate in this study.

## Author contributions

Conception and design: JR and JSK. Cohort initiation, study follow-up, data management and sample processing KoCo19 Shield: MH, MP, CP, IB, LG, NW. Cohort initiation, study follow-up, data management and sample processing PCC: HS, KA, US, JB, GI, EG, MR, CB, AP, EV. Acquisition of data: JSK, SF, VO, HS, MA. Analysis and interpretation of data: JSK and SS. Writing of the manuscript: JSK and JR. Critical reagents and manuscript editing: SF, VO, SS, and MA. Funding acquisition: MH, HS, KA, JSK, JR.

## Acknowledgements

We thank Renate Stirner and Gabriele Reiling for excellent technical assistance. The Post-COVID^LMU^ research project is financially supported by the Bavarian State Ministry for Health and Care and the Bavarian State Office for Health and Food Safety (LGL). There is a close link to the nationwide research project “Network University Medicine” (NUM), funded by the Federal Ministry of Education and Research (BMBF) (funding code: 01KX2021) and the NUM-associated research projects. JR is supported by the German Center for Infection Research (DZIF) and Else Kröner-Fresenius-Stiftung (EKFS). JSK received funding from the German Research Foundation (DFG, Deutsche Forschungsgemeinschaft), the German Diabetes Society (DDG, Deutsche Diabetes Gesellschaft), and FoeFoLe, LMU Munich. JSK is also supported by the German Society of Internal Medicine (DGIM, Deutsche Gesellschaft für Innere Medizin, Clinician Scientist Program).

## Materials and Methods

### Study design

#### KoCo19-Shield sub study of the prospective Covid19 cohort Munich -Index study (KoCo19-Index)

To establish the KoCo19-index cohort, we recruited study subjects in whose household at least one person had a PCR confirmed SARS-CoV-2 infection. Recruitment was performed as previously described^34^. In brief, individuals with PCR confirmed SARS-CoV2 infection were contacted by the responsible official authorities (City of Munich Health department) in May and June 2020. Individuals who expressed interest in participating were enrolled in the KoCo19 index study. For the KoCo19 – Shield sub study, at least one member of the participant’s household (> 14 years old) also had to be willing to participate. The KoCo19-Shield sub study comprised 177 PCR-positive individuals and 145 household members, enrolled between September 29, 2020, and January 27, 2021. Personal data of the study subjects was collected as previously described^35^. In short, the mobile data collection tool OpenDataKit (ODK) was used to capture data during study visits by field workers on Android smartphones. Study subjects completed household questionnaires as well as personal questionnaires using a web-based application. Non-responders were reminded first by email, and in case of continued non-response with a telephone reminder. Telephone interviews were offered to those who felt unable to complete the questionnaires online. Clinical demographics of study participants are reported in **Table 1**.

The study was approved by the Ethics Committee of the Medical Faculty at LMU Munich (20–275 V) and the protocol is available online (www.koco19.de)^34^. Informed consent was obtained from all enrolled participants. The study is registered to the German Clinical Trials Register (DRKS-ID: DRKS00022155).

#### Post Covid Study

Post-COVID-Care (PCC) study is an ongoing prospective single-center study comprised of patients with persistent symptoms following acute COVID-19. Participants with COVID-19 sequelae were recruited from the Post-COVID outpatient clinic at the University Hospital LMU Munich. Samples were collected from participants enrolled between April and July 2022. Inclusion criteria were age ≥ 18 years; persistent symptoms > 12 weeks within 6 months following initial COVID-19 infection. Pre-specified exclusion criteria were other explanations for the symptom onset or complete resolution of symptoms. All participants were scheduled for follow-up for at least 6 months and up to 24 months if symptoms persisted. At baseline and during the routine follow-up visits blood samples were obtained and each patient completed progressive web app (PWA)-based questionnaires (LCARS-C, LMU Munich). Patients who did not undergo any follow-up on site were asked to fill out the follow-up surveys using the PWA-based questionnaire at home using a PC, smartphone or tablet. Informed consent was obtained from all participants before inclusion into the study. Clinical characteristics of study participants are reported in **Table 1**. The study was approved by the Ethics Committee of the Medical Faculty at LMU Munich (No. 21-1165) and registered to the German Clinical Trials Register (DRKS-ID: DRKS00030974).

#### Participant Surveys

KoCo19 and PCC study participants completed a comprehensive suite of surveys, combining validated patient-reported outcomes (PROs) with custom, purpose-developed tools by the study team. Baseline demographic data collected from surveys included gender, age, body mass index (BMI), race, and medical comorbidities. Additionally, participants in the Long COVID and convalescent group were asked to provide COVID-19 clinical data including date of symptom onset and acute disease severity (non-hospitalized vs. hospitalized), SARS-CoV-2 polymerase chain reaction (PCR) diagnostic testing results, and SARS-CoV-2 antibody testing results. All participants were asked to report SARS-CoV-2 vaccination status including date of vaccinations and vaccine brand.

#### Blood sample processing

Whole blood was collected in four potassium-EDTA-coated blood collection tubes (Sarstedt) from participants at University Hospital, LMU, Munich, Germany. Following blood draw, all participant samples were assigned unique study identifiers and de-identified by research staff. Blood samples were processed the same day as collection. Plasma samples were collected after centrifugation of whole blood at 450×g for 10 minutes at room temperature (RT). Plasma was then transferred to 1,8-ml polyethylene Cryotube™ vials (ThermoFisher), aliquoted, and stored at -80°C. For isolation of peripheral blood mononuclear cells (PBMCs), two tubes each of the remaining whole blood sample were pooled and filled up to a total volume of 32.5 ml with Hank’s Balanced Salts Solution (Capricorn or Sigma). 13,5ml Histopaque®-1077 (Sigma) were added at the bottom of each tube and samples were centrifuged at 450xg for 30 minutes at RT without break. PBMC layer on top of the Histopaque® layer was collected and washed twice in Hank’s balanced salts solution. Isolated cells were counted using a CASY cell counter and analyzer (Schärfe System GmbH) before storage in liquid nitrogen at -180°C for cryopreservation.

#### Flow cytometry

Cryopreserved PBMCs were thawed in a 37°C water bath, pipetted into Iscove’s Modified Dulbecco’sMedium (IMDM) supplemented with 10% FCS medium, and washed by centrifugation. Three to six million cells per sample were incubated with antibodies to surface antigens (Table S1) for 30 minutes at 4°C, washed with FACS buffer (1XDPBS, 3% FCS, 0.05% NaN3), fixed with 2% paraformaldehyde for 10 minutes, washed again with FACS buffer, and resuspended in FACS buffer. Flow cytometry was performed on BD LSRFortessa X-20. Fluorochrome compensation was performed with single-stained UltraComp eBeads (Invitrogen, Cat# 01-2222-42). Samples were FSC-A/SSC-A gated to exclude debris, followed by FSC-H/FSC-A gating to select single cells and Zombie NIR fixable or DAPI to exclude dead cells. Innate lymphoid cells were identified as lineage negative (CD1a^-^, CD14^-^, CD19^-^, CD34^-^, CD94^-^, CD123^-^, FcER1a^-^, TCRab^-^, TCRgd^-^, BDCA2^-^), CD45^+^, CD161^+^, CD127^+^, as indicated. The full gating strategy is shown in Fig. S1 and was adapted from previous work^29^. Data were analyzed using FlowJo version 10.7 software (TreeStar, USA) and compiled using Prism (GraphPad Software). T-distributed stochastic neighbor embedding (t-SNE) visualization of flow cytometry data was performed using Cytobank.

#### Quantification of plasma cytokine levels

The plasma levels of 46 molecular species were quantified using a Luminex platform (Human Cytokine Discovery, R&D System, Minneapolis, MN) for the simultaneous detection of the following molecules: G-CSF, PDGF-AA, EGF, PDGF-AB/BB, VEGF, GM-CSF, FGF, GRZB, IL-1A, IL-1RA, IL-2, IL-27, IL-4, IL-6, IL-10, IL-13, TNF, IL-17C, IL-11, IL-18, IL-23, IL-6RA, IL-19, IFN-B, IL-3, IL-5, IL-7, IL-12p70, IL-15, IL-33, TGF-B, IFN-G, IL-1B, IL-17, IL-17E, CCL3, CCL11, CCL20, CXCCL1, CXCL2, CCL5, CCL2, CCL4, CCL19, CXCL1, CXCL10, PD-L1, FLT-3, TACI, FAS, LEPTIN R, APRIL, OPN, BAFF, LEPTIN, BMP4, CD40 LIGAND, FAS LIGAND, BMP7, BMP2, and TRAIL, according to the manufacturer’s instruction

#### Statistical analysis

All data are expressed as means ± standard error of the mean (SEM) unless otherwise noted. Comparisons between two groups were analyzed by using unpaired two-tailed Student’s t-tests, and multiple comparisons were analyzed by one-way analysis of variance (ANOVA) with Tukey’s multiple comparisons test (Prism, GraphPad Software, La Jolla, CA), with * = p<0.05, ** = p<0.01, *** =p<0.001, **** =p<0.0001. Each symbol reflects individuals for flow analysis or plasma cytokine levels.

#### Unsupervised data analysis

Cytobank^36^ was used for initial manual gating of Lineage-negative cells and ILC subsets ILC1, ILC2, and ILCP, using the same gating strategy as described above. FCS files were transformed with. Lineage-negative cells were subjected to dimensionality reduction using Cytobank opt-SNE with default hyperparameters and following embedding markers with normalized scales Cytobank arcsinh transformation: CD117, CD127, CD161, CD45RA, CD56, CRTH2, HLA-DR, and SLAMF1. All pre-gated events were used without prior downsampling from 91 samples. In order to perform downstream statistical analyses in R (http://www.r-project.org/) and visualize t-SNE maps across the 91 samples, events within ILC subsets were exported from Cytobank as tab-separated values containing compensated and transformed marker expression levels as well as t-SNE coordinates and metacluster assignment. T-SNE plots were generated after subsampling each sample to contain a maximum of 2500 events. High-resolution group differences were visualized by calculating Cohen’s D for a given comparison across the t-SNE map. To this end, we generated adaptive 2D histograms using the probability binning algorithm available through the R *flowFP* package^37^. Dependent on the total number of cells available, a single binning model was created on collapsed data from all samples, by recursively splitting the events at the median values along the two t-SNE dimensions. We chose a grid of 256 bins to have, on average, at least eight cells per bin in each sample for statistical accuracy. Since there was a significant difference between cellular frequency distributions between the six measurement days, the batch effect was first regressed out by fitting a linar model to each bin after applying the arcsine-square-root transformation for proportions. The group-difference effect sizes were then calculated for each bin using the cohen.d function of the *effsize* package. In order to get a smoothed representation of the effect size map, adaptive binning was performed on a series of rotated coordinates and per cell-averaged effect size values were used to color-encode each cell throughout the t-SNE map. All analyses were performed using R version 4.1.1, available free online at https://www.r-project.org.

### Post Covid Care group members

Kristina Adorjan, Shahnaz C. Azad, Petra Bäumler, Christopher Bensch, Johannes Bogner, Svenja Anike Feldmann, Fides Heimkes, Gerardo Ibarra, Dominik Irnich, Anna-Lena Johlke, Stefan Kääb, Kathrin Kahnert, Eduard Kraft, Katrin Milger-Kneidinger, Veronica Ober, Anna Pernpruner, Jan Rémi, Julia Roider, Michael Ruzicka, Simone Sachenbacher, Florian Schöberl, Konstantin Stark, Andreas Straube, Hans Christian Stubbe, Elisabeth Valdinoci, Martin Weigl, Nora Wunderlich.

### KoCo19 study group members

Emad Alamoudi, Jared Anderson, Abhishek Bakuli, Marc Becker, Franziska Bednarzki, Olimbek Bemirayev, Jessica Beyerl, Patrick Bitzer, Rebecca Boehnlein, Friedrich Caroli, Lorenzo Contento, Alina Czwienzek, Flora Deák, Maximilian N. Diefenbach, Gerhard Dobler, Jürgen Durner, Judith Eckstein, Philine Falk, Volker Fingerle, Felix Forster, Turid Frahnow, Guenter Froeschl, Otto Geisenberger, Kristina Gillig, Philipp Girl, Pablo Gutierrez, Anselm Haderer, Marlene Hannes, Jan Hasenauer, Tim Haselwarter, Alejandra Hernandes, Matthias Herrmann, Leah Hillari, Christian Hinske, Tim Hofberger, Sacha Horn, Kristina Huber, Christian Janke, Ursula Kappl, Antonia Kessler, Zohaib N. Khan, Johanna Kresin, Arne Kroidl, Magdalena Lang, Silvan Lange, Michael Laxy, Ronan Le Gleut, Reiner Leidl, Leopold Liedl, Xhovana Lucaj, Petra Mang, Alisa Markgraf, Rebecca Mayrhofer, Dafni Metaxa, Hannah Mueller, Katharina Mueller, Laura Olbrich, Ivana Paunovic, Claire Pleimelding, Michel Pletschette, Stephan Prueckner, Kerstin Puchinger, Peter Puetz, Katja Radon, Elba Raimundéz, Jakob Reich, Friedrich Riess, Camilla Rothe, Viktoria Ruci, Nicole Schaefer, Yannik Schaelte, Benedikt Schluse, Elmar Saathoff, Lara Schneider, Mirjam Schunk, Lars Schwettmann, Peter Sothmann, Kathrin Strobl, Jeni Tang, Fabian Theis, Verena Thiel, Jonathan von Lovenberg, Julia Waibel, Claudia Wallrauch, Roman Woelfel, Julia Wolff, Tobias Wuerfel, Sabine Zange, Eleftheria Zeggini, Anna Zielke.

## References

1. Clark D V, Kibuuka H, Millard M, et al. Long-term sequelae after Ebola virus disease in Bundibugyo, Uganda: a retrospective cohort study. Lancet Infect Dis. 2015;15(8):905–912. doi:10.1016/S1473-3099(15)70152-0

2. A Post-Graduate Lecture ON THE NERVOUS SEQUELÆ OF INFLUENZA. Lancet. 1893;142(3645):73–76. doi:https://doi.org/10.1016/S0140-6736(00)65088-2

3. Fluge Ø, Tronstad KJ, Mella O. Pathomechanisms and possible interventions in myalgic encephalomyelitis/chronic fatigue syndrome (ME/CFS). J Clin Invest. 2021;131(14). doi:10.1172/JCI150377

4. Newton DJ, Kennedy G, Chan KKF, Lang CC, Belch JJF, Khan F. Large and small artery endothelial dysfunction in chronic fatigue syndrome. Int J Cardiol. 2012;154(3):335–336. doi:10.1016/j.ijcard.2011.10.030

5. Datta SD, Talwar A, Lee JT. A Proposed Framework and Timeline of the Spectrum of Disease Due to SARS-CoV-2 Infection: Illness Beyond Acute Infection and Public Health Implications. JAMA. 2020;324(22):2251–2252. doi:10.1001/jama.2020.22717

6. Sivan M, Taylor S. NICE guideline on long covid. BMJ. 2020;371:m4938. doi:10.1136/bmj.m4938

7. Crook H, Raza S, Nowell J, Young M, Edison P. Long covid—mechanisms, risk factors, and management. BMJ. 2021;374:n1648. doi:10.1136/bmj.n1648

8. Margalit I, Yelin D, Sagi M, et al. Risk Factors and Multidimensional Assessment of Long Coronavirus Disease Fatigue: A Nested Case-Control Study. Clin Infect Dis. 2022;75(10):1688–1697. doi:10.1093/cid/ciac283

9. Choutka J, Jansari V, Hornig M, Iwasaki A. Unexplained post-acute infection syndromes. Nat Med. 2022;28(5):911–923. doi:10.1038/s41591-022-01810-6

10. Kreutmair S, Unger S, Núñez NG, et al. Distinct immunological signatures discriminate severe COVID-19 from non-SARS-CoV-2-driven critical pneumonia. Immunity. 2021;54(7):1578–1593.e5. doi:10.1016/j.immuni.2021.05.002

11. Bergamaschi L, Mescia F, Turner L, et al. Longitudinal analysis reveals that delayed bystander CD8+ T&#xa0;cell activation and early immune pathology distinguish severe COVID-19 from mild disease. Immunity. 2021;54(6):1257–1275.e8. doi:10.1016/j.immuni.2021.05.010

12. Blot M, Bour J-B, Quenot JP, et al. The dysregulated innate immune response in severe COVID-19 pneumonia that could drive poorer outcome. J Transl Med. 2020;18(1):457. doi:10.1186/s12967-020-02646-9

13. Lim J, Puan KJ, Wang LW, et al. Data-Driven Analysis of COVID-19 Reveals Persistent Immune Abnormalities in Convalescent Severe Individuals. Front Immunol. 2021;12. https://www.frontiersin.org/articles/10.3389/fimmu.2021.710217

14. Varga Z, Flammer AJ, Steiger P, et al. Endothelial cell infection and endotheliitis in COVID-19. Lancet. 2020;395(10234):1417–1418. doi:10.1016/S0140-6736(20)30937-5

15. Cheon IS, Li C, Son YM, et al. Immune signatures underlying post-acute COVID-19 lung sequelae. Sci Immunol. 2022;6(65):eabk1741. doi:10.1126/sciimmunol.abk1741

16. Klein J, Wood J, Jaycox J, et al. Distinguishing features of Long COVID identified through immune profiling. medRxiv. Published online January 2022:2022.08.09.22278592. doi:10.1101/2022.08.09.22278592

17. Phetsouphanh C, Darley DR, Wilson DB, et al. Immunological dysfunction persists for 8 months following initial mild-to-moderate SARS-CoV-2 infection. Nat Immunol. 2022;23(2):210–216. doi:10.1038/s41590-021-01113-x

18. Moro K, Yamada T, Tanabe M, et al. Innate production of TH2 cytokines by adipose tissue-associated c-Kit+Sca-1+ lymphoid cells. Nature. 2010;463(7280):540–544. doi:10.1038/nature08636

19. Vivier E, Artis D, Colonna M, et al. Innate Lymphoid Cells: 10 Years On. Cell. 2018;174(5):1054–1066. doi:10.1016/j.cell.2018.07.017

20. Cautivo KM, Matatia PR, Lizama CO, et al. Interferon gamma constrains type 2 lymphocyte niche boundaries during mixed inflammation. Immunity. 2022;55(2):254–271.e7. doi:10.1016/j.immuni.2021.12.014

21. Dahlgren MW, Molofsky AB. All along the watchtower: group 2 innate lymphoid cells in allergic responses. Curr Opin Immunol. 2018;54:13–19.

22. Gomez-Cadena A, Spehner L, Kroemer M, et al. Severe COVID-19 patients exhibit an ILC2 NKG2D+ population in their impaired ILC compartment. Cell Mol Immunol. 2021;18(2):484–486. doi:10.1038/s41423-020-00596-2

23. Silverstein NJ, Wang Y, Manickas-Hill Z, et al. Innate lymphoid cells and COVID-19 severity in SARS-CoV-2 infection. Giamarellos-Bourboulis EJ, Rath S, Giamarellos-Bourboulis EJ, Kyriazopoulou E, eds. Elife. 2022;11:e74681. doi:10.7554/eLife.74681

24. Antonelli M, Penfold RS, Merino J, et al. Risk factors and disease profile of post-vaccination SARS-CoV-2 infection in UK users of the COVID Symptom Study app: a prospective, community-based, nested, case-control study. Lancet Infect Dis. 2022;22(1):43–55. doi:10.1016/S1473-3099(21)00460-6

25. Magnusson K, Kristoffersen DT, Dell’Isola A, et al. Post-covid medical complaints following infection with SARS-CoV-2 Omicron vs Delta variants. Nat Commun. 2022;13(1):7363. doi:10.1038/s41467-022-35240-2

26. Kaiser R, Leunig A, Pekayvaz K, et al. Self-sustaining IL-8 loops drive a prothrombotic neutrophil phenotype in severe COVID-19. JCI Insight. 2021;6(18). doi:10.1172/jci.insight.150862

27. Zhang Z, Ai G, Chen L, et al. Associations of immunological features with COVID-19 severity: a systematic review and meta-analysis. BMC Infect Dis. 2021;21(1):738. doi:10.1186/s12879-021-06457-1

28. Sabbatino F, Conti V, Franci G, et al. PD-L1 Dysregulation in COVID-19 Patients. Front Immunol. 2021;12. https://www.frontiersin.org/articles/10.3389/fimmu.2021.695242

29. Mazzurana L, Bonfiglio F, Forkel M, D’Amato M, Halfvarson J, Mjösberg J. Crohn’s Disease Is Associated With Activation of Circulating Innate Lymphoid Cells. Inflamm Bowel Dis. 2021;27(7):1128–1138. doi:10.1093/ibd/izaa316

30. Lim AI, Li Y, Lopez-Lastra S, et al. Systemic Human ILC Precursors Provide a Substrate for Tissue ILC Differentiation. Cell. 2017;168(6):1086–1100.e10. doi:10.1016/j.cell.2017.02.021

31. Kokkinou E, Pandey RV, Mazzurana L, et al. CD45RA+CD62L− ILCs in human tissues represent a quiescent local reservoir for the generation of differentiated ILCs. Sci Immunol. 2023;7(70):eabj8301. doi:10.1126/sciimmunol.abj8301

32. Fonseca W, Lukacs NW, Elesela S, Malinczak C-A. Role of ILC2 in Viral-Induced Lung Pathogenesis. Front Immunol. 2021;12.

33. Vanoni G, Ercolano G, Candiani S, et al. Human primed ILCPs support endothelial activation through NF-κB signaling. Cerullo V, Taniguchi T, eds. Elife. 2021;10:e58838. doi:10.7554/eLife.58838

34. Brand I, Gilberg L, Bruger J, et al. Broad T Cell Targeting of Structural Proteins After SARS-CoV-2 Infection: High Throughput Assessment of T Cell Reactivity Using an Automated Interferon Gamma Release Assay. Front Immunol. 2021;12.

35. Pritsch M, Radon K, Bakuli A, et al. Prevalence and Risk Factors of Infection in the Representative COVID-19 Cohort Munich. Int J Environ Res Public Health. 2021;18(7). doi:10.3390/ijerph18073572

36. Kotecha N, Krutzik PO, Irish JM. Web-based analysis and publication of flow cytometry experiments. Curr Protoc Cytom. 2010;Chapter 10:Unit10.17-Unit10.17. doi:10.1002/0471142956.cy1017s53

37. Rogers WT, Holyst HA. FlowFP: A Bioconductor Package for Fingerprinting Flow Cytometric Data. Gottardo R, ed. Adv Bioinformatics. 2009;2009:193947. doi:10.1155/2009/193947

