## Supplementary Information for "Persistent immune abnormalities discriminate post-COVID syndrome from convalescence"

#### Supplementary Material

##### Supplementary Figure 1

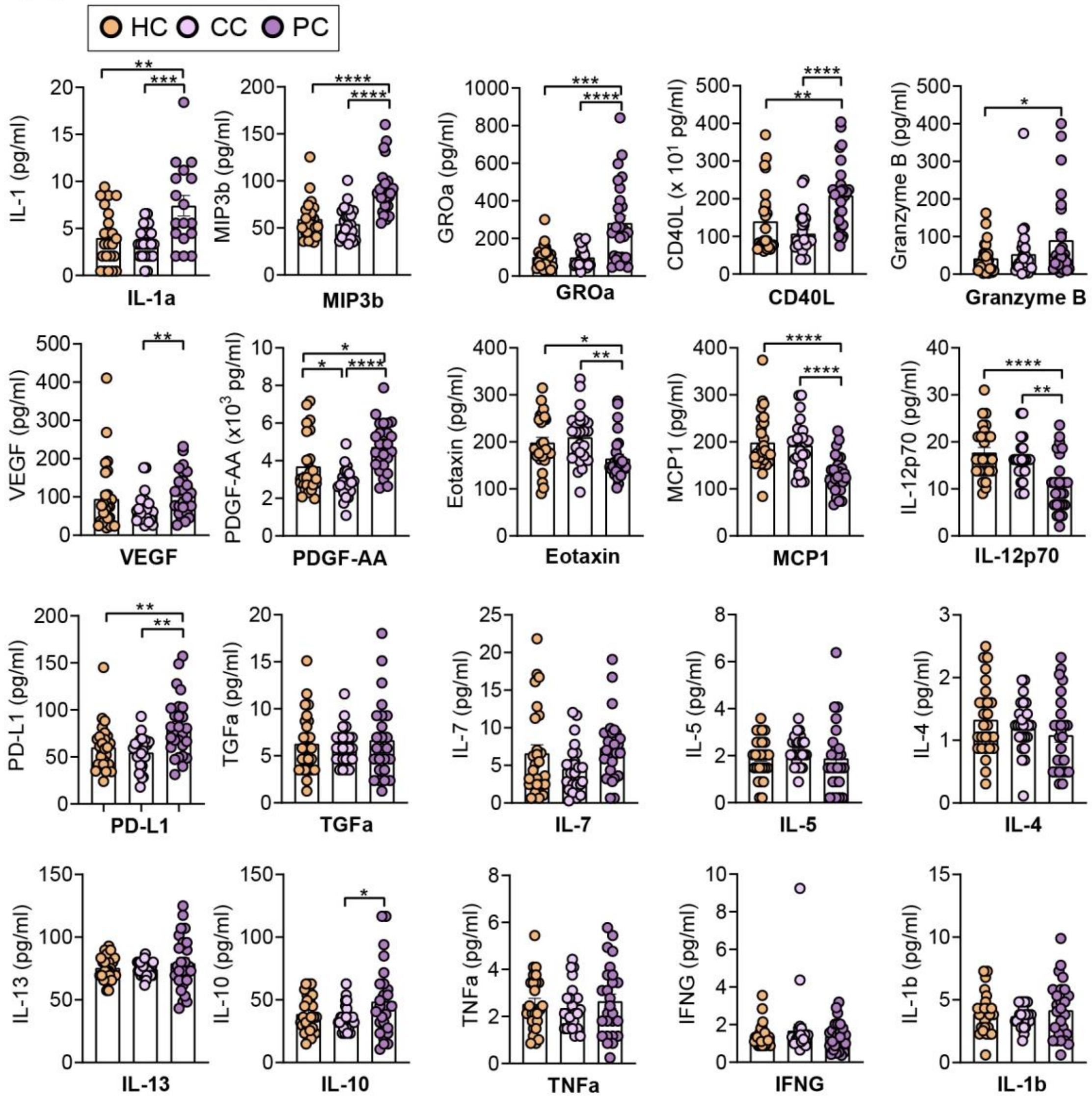


**Supplementary Figure 1: Post-COVID participants display altered plasma cytokine expression levels.**

Multiplex assay quantification showing levels of IL-1a, MIP3b, CXCL1/Groa, CD40L, Granzyme B, VEGF, PDGF-AA, Eotaxin, MCP1, IL-12p70, PDL-1, TGFa, IL-7, IL-5, IL-4, IL-13, IL-10, TNFa, IFNG, and IL-1b in plasma of healthy controls with no prior SARS-CoV-2 infection (HC), n=32, convalescent SARS-CoV-2 participants without persisting symptoms (CC), n=32, and convalescent SARS-CoV-2 participants with persisting symptoms (PC), n=27 at 3-10 months after acute COVID infection.

Bar graphs indicate mean (±SE), unpaired t-tests, *p ≤ 0.05, **p ≤ 0.01, ***p ≤ 0.001, ****p ≤ 0.0001.

### Supplementary Figure 2

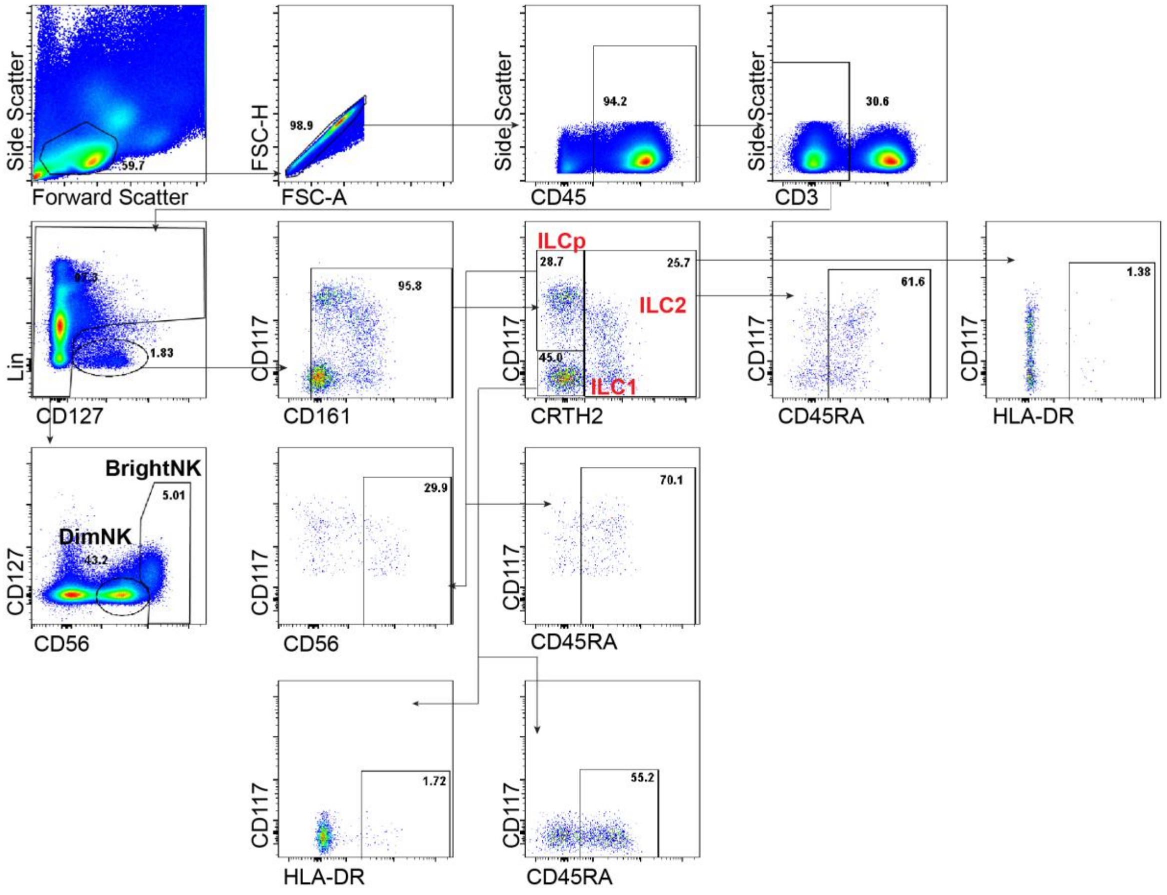


**Supplementary Figure 2: Flow cytometry gating scheme.**

Representative flow cytometry gating scheme for analysis of innate lymphoid cells (ILCs) in peripheral blood mononuclear cells (PBMCs). Numbers represent percentages of cells in respective gates. Lineage markers contained CD1a, CD14, CD19, CD123, BDCA2, FceR1, CD34, CD94, TCRαβ, TCRγδ, FceR1, and dead cell marker.

### Supplementary Figure 3

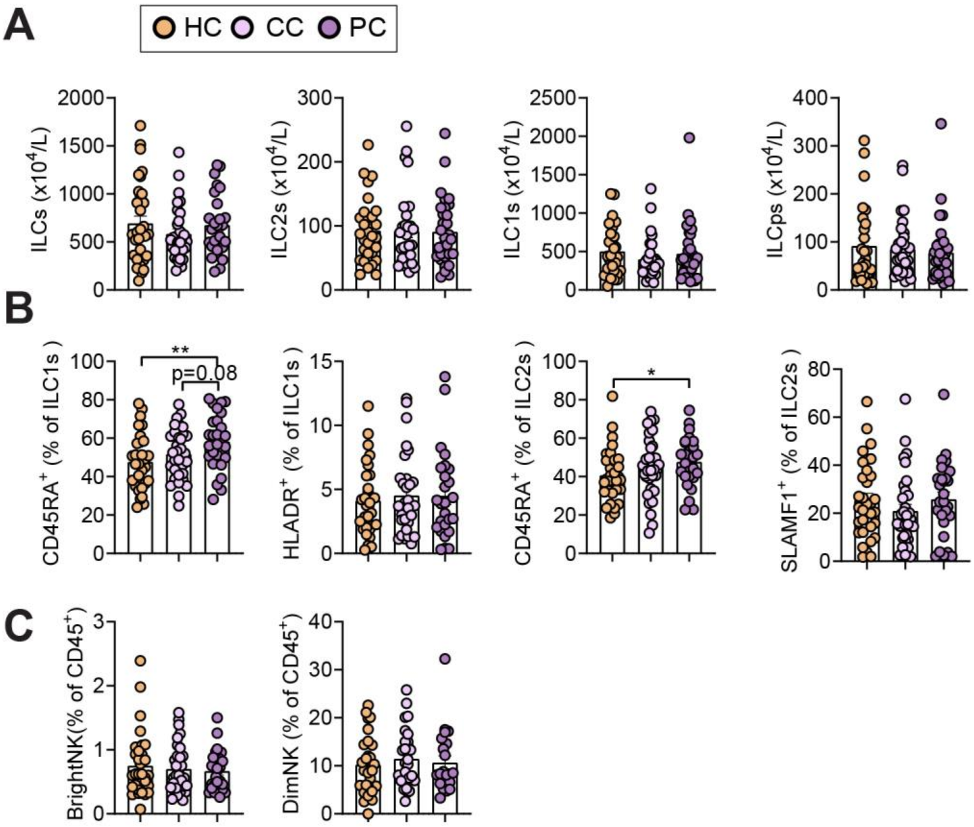


**Supplementary Figure 3: Levels of innate lymphoid cells among study groups.** (A-C) Flow cytometry quantification, showing total numbers (A) and percent (B, C) of innate lymphoid cells (ILCs) and NK cells in peripheral blood mononuclear cells (PBMCs) of healthy controls with no prior SARS-CoV-2 infection (HC), convalescent SARS-CoV-2 participants without persisting symptoms (CC) and convalescent SARS-CoV-2 participants with persisting symptoms (PC) at 3-10 months after acute COVID infection.

Bar graphs indicate mean (±SE), unpaired t-tests (A-C), $*p \leq 0.05$.
